# Safety and Efficacy of Cardiac Sympathetic Denervation for Ventricular Arrhythmias: An Updated Review and Meta-Analysis

**DOI:** 10.1101/2024.08.09.24311768

**Authors:** Daniel B. Hanna, Ahmadreza Karimianpour, Nicole Mamprejew, Chris Fiechter, Dhiran Verghese, Juan Sierra, Viviana Navas, Dinesh Sharma

## Abstract

**Introduction:** The role of sympathetic nervous system in the initiation and continuation of ventricular tachyarrhythmias (VTA) is well established. However, whether CSD reduces implantable cardioverter defibrillator (ICD) shocks and recurrent VTA is still uncertain.

**Objective:** To explore long-term arrhythmic outcomes, safety, and efficacy of CSD by measuring event rates of recurrent VTA and ICD shocks after CSD.

**Methods:** A comprehensive literature search was performed at Medline and Embase until March 2023. Our primary outcome was event rate of ICD shocks and VTA. Analyses were conducted using Comprehensive Meta-Analysis software.

**Results:** Initial search yielded 1,324 scientific studies with a total of 15 studies fitting our inclusion criteria. ICD shocks at 1 year post CSD revealed an event rate of 69.8% (95% CI, 56.4% – 80.4% with 50% heterogeneity) (I^2^ statistic)). ICD shocks at 6 months had an event rate of 59.1% (95% CI 46.9% - 70.4%, 47 I^2^). Freedom of VTA 1 year post CSD revealed an event rate of 64.3% (95% CI, 42.3% - 81.5%, 26% I^2^). Freedom from VTA at 6 months revealed an event rate of of 62.3% (95% CI, 51.2% - 72.2%, 40% I^2^). Reported mortality due to VTA was subdivided into short-term (0-30 days), intermediate-term (31-364 days) and long-term (32-364 days). The event rate for the short-term tertile was 8.9% (95% CI, 5.0% - 15.4%, 0% I^2^), medium-term was 5.3% (95% CI, 2.4% - 11.3%, 0% I^2^) and long-term 5.2% (95% CI, 2.4% - 10.9%, 0% I^2^).

**Conclusion:** CSD seems to be promising as an acceptable treatment strategy for recurrent VTA refractory to traditional pharmacological or ablation therapy.

## Introduction

The annual incidence of sudden cardiac death (SCD) ranges from 180,000–300,000 in the United States (US) and 4,250,000 worldwide.^1^ Moreover, 25% of all deaths is due to SCD in the form of ventricular tachycardia arrhythmias (VTA)^2^. In patients protected from SCD with an implantable cardioverter defibrillator (ICD), recurrent VTA and ICD therapies increases mortality^2^. The autonomic nervous system (ANS) is a key component in the development of recurrent VTA. The interplay between sympathetic and parasympathetic inputs exert profound effects on cardiac electrophysiology and predispose individuals to the development and worsening of arrhythmias, notably VTA. ^3^

Increased sympathetic tone augments automaticity and alters myocardial conduction properties^3^. These findings have prompted investigation of sympathetic modulation strategies as a means to manage refractory ventricular arrhythmias.

Cardiac sympathetic denervation (CSD) has emerged as a promising intervention. The primary objective is to reduce sympathetic tone and thus arrhythmia burden to help improve long-term mortality and quality of life. This method was originally explored to address VTA in inherited channelopathies such as catecholaminergic polymorphic ventricular tachycardia, however, CSD appears to be a potentially viable treatment option for patients with VTA in the setting of structural heart disease.^4,5^ CSD has demonstrated efficacy in reducing VTA storm refractory to conventional antiarrhythmic drug therapy and catheter ablation, which highlights its potential as a therapeutic strategy in patients hospitalized with VTA storm.^6^

The utility of CSD extends beyond ischemic cardiomyopathies and may also prove useful in treatment of multimorphology VTA in nonischemic cardiomyopathies (NICM) and where multifocal premature ventricular contractions present a significant challenge to standard catheter ablation techniques.^7^ In these cases, CSD may be an effective alternative for arrhythmia control and symptom management. There is limited data regarding the long-term efficacy and safety of CSD in patients. Specific to HF, further investigation is warranted to determine the broader positive or negative physiologic effects other than suppressing arrhythmias.^8^ Thus, the aim of this contemporary systematic review and meta-analysis is to provide updated insights into the safety and efficacy of CSD regarding patients with structural heart disease suffering from recurrent VTA.

## Methods

A comprehensive literature search was performed at Medline and Embase from inception until March 2023. The search terms “Sympathectomy” OR “Cardiac sympathetic denervation” OR “Refractory ventricular tachycardia” OR “Cardiac Denervation” OR “Electrical storm” OR “Ventricular arrhythmia,” were used. A total of four investigators agreed on the final study selection and independently searched the complete database. The inclusion criteria included any study which reported the short- or long-term outcomes of CSD in the management of VTA if a) age group was >18 years of age b) mortality was reported c) there was an occurrence of VTA or electrical storm, or d) follow-up data for at least 1 month. Individual case reports were not included. Data obtained included study design, follow-up duration, number of patients, baseline demographics, comorbidities, type of ventricular arrhythmia, type of cardiomyopathy, and our primary and secondary outcomes. This was compiled through PRISMA (Preferred Reporting Items for Systematic Reviews and Meta-Analyses (Figure 1).

**Figure 1.**
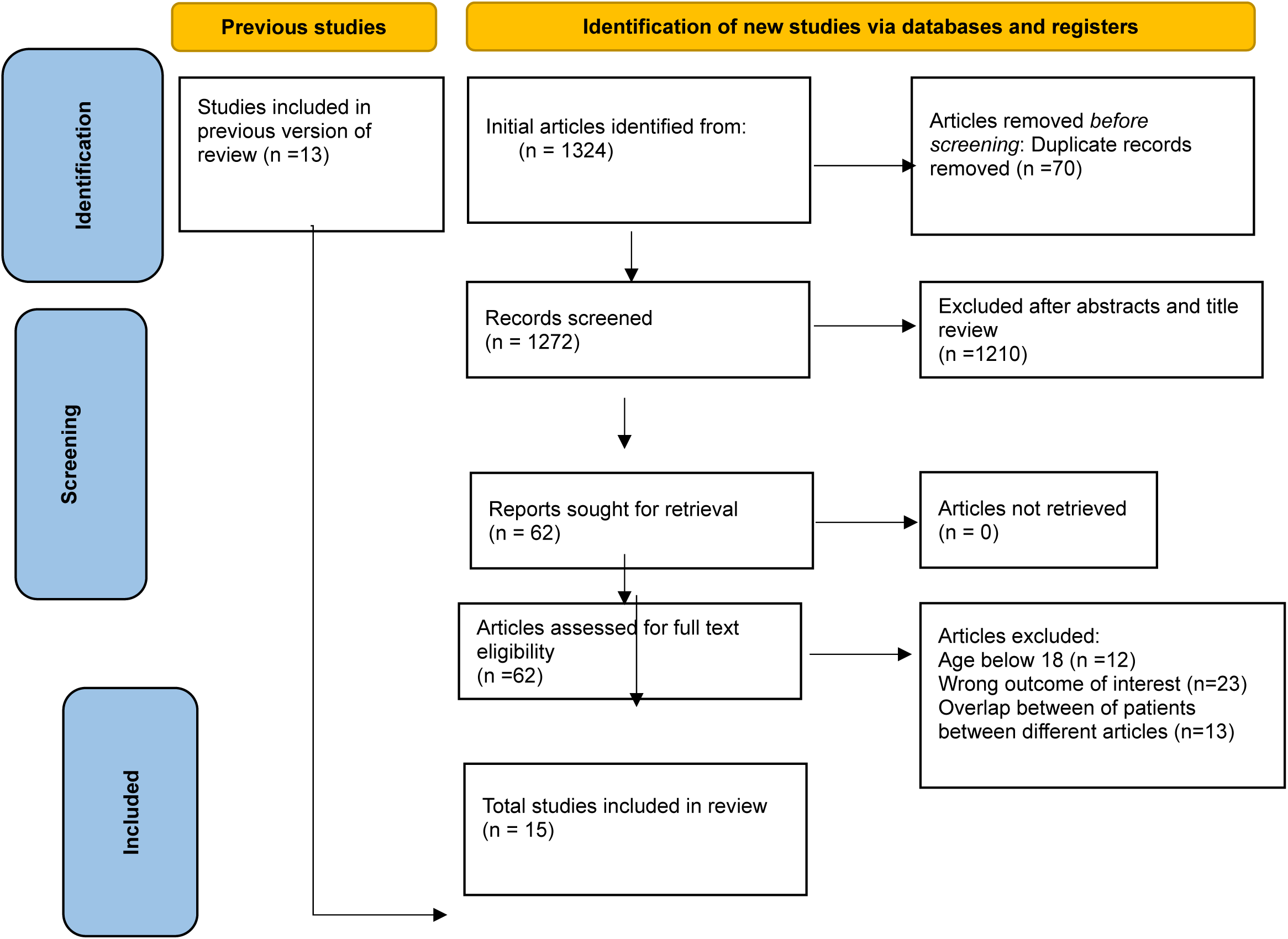
PRISMA Analysis

### Statistical analysis

Statistical analysis was conducted using a single-arm-meta-analysis model using CMA software (Comprehensive Meta-Analysis Version 4.0) with a random-effects model to calculate overall event rate. Study heterogeneity was evaluated with Cochran’s Q and I2 index.

## Results

The initial search yielded 1,324 scientific studies, compiled in PRISMA for inclusion, exclusion and analysis as shown in Figure 1. After the removal of duplicates, 1272 underwent title and abstract review as primary survey which excluded 1,210 studies. The remaining 62 studies underwent full-text review and 38 were excluded due to a lack of reporting outcomes of interests, overlap of patients or insufficient data. This yielded 15 studies with a total of 373 patients that met inclusion our criteria^9–22^. Using the Newcastle-Ottawa quality assessment form, thirteen were identified as good quality while two were of fair quality. Baseline characteristics of the included studies are listed in Table 1.

**Table 1:** Baseline Characteristics of all studies.

| Author's Last Name, Year | Journal | Sample Size | Left Sided CSD | Right Sided CSD | Bilateral CSD | Mean Average Follow up (days) | Mean Age in years | Average Ejection Fraction (%) | Study Type |
| --- | --- | --- | --- | --- | --- | --- | --- | --- | --- |
| Barwad, 2021 | J Cardiovasc Electrophysio | 65 | 5 | 0 | 60 | 821.2 ± 730 | 50 ± 18 | 28 | Retrospect ive |
| Cai, 2019 | J Cardiovasc Electrophysio | 19 | 14 | 0 | 5 | 1064 ± 811 | 54.5 ± 16.6 | 43.6 ± 18.3 | Prospectiv e |
| Cardona-Guarache, 2017 | J Cardiovasc Electrophysio | 6 | 0 | 0 | 6 | N/A | 55.1 | 30.8 | Case Series |
| Juhos, | Bratisl Med J | 3 | 0 | 0 | 3 | N/A | 42.6 | N/A | Case |

| 2020 |  |  |  |  |  |  |  |  | Series |
| --- | --- | --- | --- | --- | --- | --- | --- | --- | --- |
| **König, 2022** | ESC | 21 | 2 | 0 | 19 | 276.79<br>2 | 63.7+14<br>.4 | 32.5+10.9 | Retrospect<br>ive |
| **Krause, 2022** | Ann Thorac Surg | 12 | 9 | 0 | 3 | 358 | 63.7 +<br>8.9 | 20 | Retrospect<br>ive |
| **Lee, 2022** | Interactive CardioVascular and Thoracic Surgery | 13 | 0 | 0 | 13 | 264.62<br>5 | 66 | 41.66 | Retrospect<br>ive |
| **Melinosky, 2023** | Innovations: Technology and Techniques in Cardiothoracic and Vascular Surgery | 67 | 2 | 2 | 63 | 1163 | 54 | N/A | Retrospect<br>ive |
| **Miki, 2022** | Int Heart J | 4 | 0 | 0 | 4 | 119.5 | 55 ± 22 | 30.0 ± 8.2 | Retrospect<br>ive |
| **Park, 2023** | J Chest Surg | 10 | 9 | 0 | 0 | 535.33 | 54 | 58 | Retrospect<br>ive |
| **Prabhu, 2016** | J Interv Card Electrophysiology | 9 | 0 | 0 | 10 | 1003.7<br>5 | 52 | 33 | Retrospect<br>ive |
| **Richardson, 2017** | Heart Rhythm | 7 | 1 | 0 | 6 | N/A | 50.8 | N/A | Case<br>Series |
| **Vaseghi 2017** | JACC | 121 | 23 | 0 | 98 | 547.5 | 55 | 30 | Retrospecti<br>ve<br>multicenter<br>registry |
| **Velásquez 2019** | Arch Cardiol Mex | 6 | 6 | 0 | 0 | 730 | 56 | N/A | Retrospect<br>ive |
| **Yalin, 2020** | Clinical Research in Cardiology | 10 | 6 | 0 | 4 | 304 | 61.6 ±<br>19.6 | 41.66 | Retrospect<br>ive |

Our final 15 studies yielded a total of 373 patients with a mean age of 55 ± 5.3 years with 72% ± 24% being male and a mean follow-up of 477.8 ± 317.7 months. Average left ventricular ejection fraction (EF) amongst all studies was found to be 30.5% ± 9.2% with the most common type of heart failure being ischemic cardiomyopathy with a prevalence of 31%. Demographic of patients can be found in Table 2. The most common type of ventricular arrhythmia was monomorphic VT (17%) (Table 3). Of the 15 studies, 14 included surgical complications. We found that both neuropathic pain and hypotension was the most common complications with an incidence of 20.5% (Figure 2). Of the total 373 patients 79% (n=291) underwent bilateral, 21% (n=77) left sided, and 0.5% (n=2) right sided CSD. All of the patients underwent CSD using video-assisted thoracoscopic surgery (VATS).

**Figure 2.**
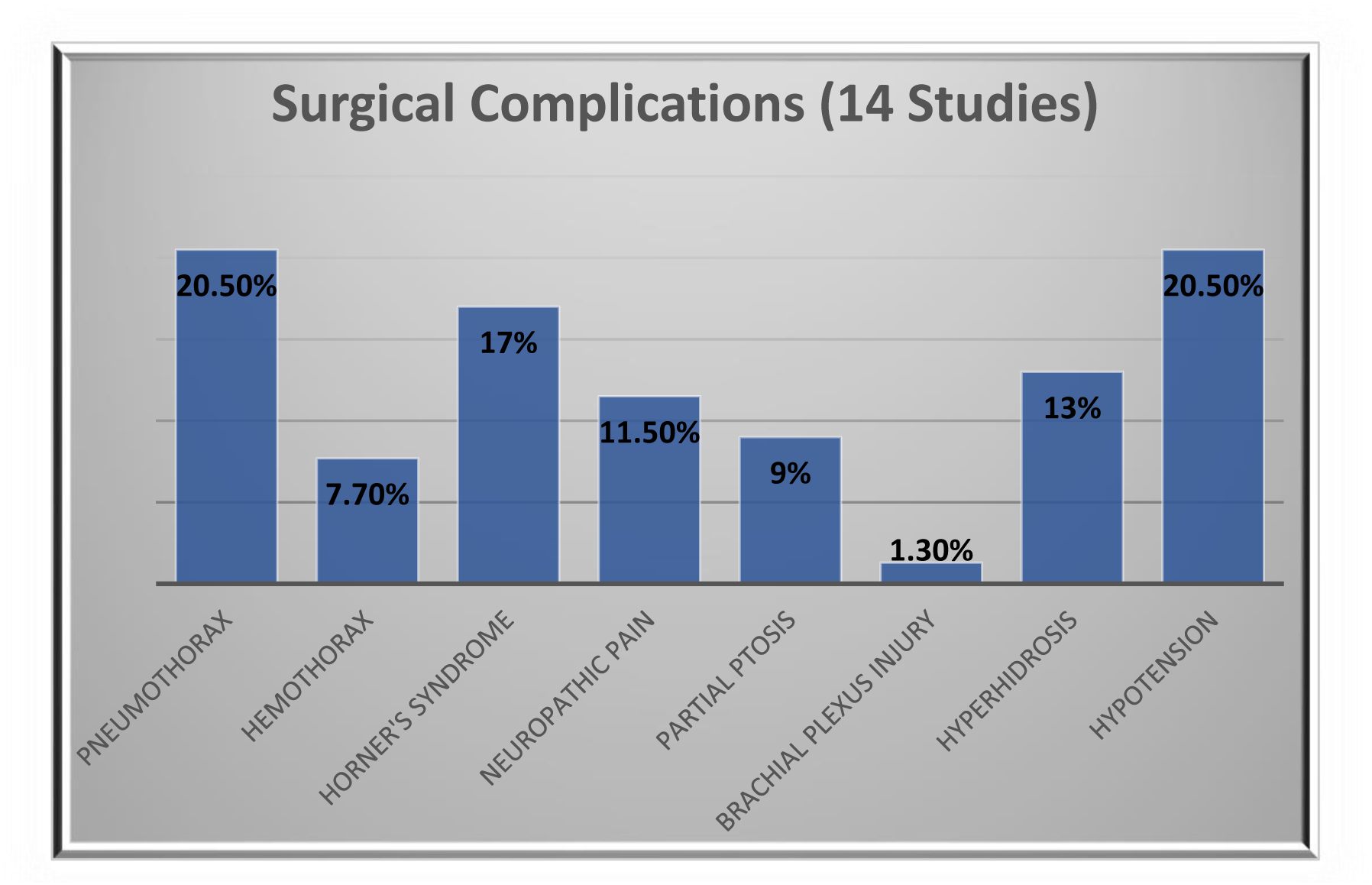
Post surgical complications.

**Table 2.**
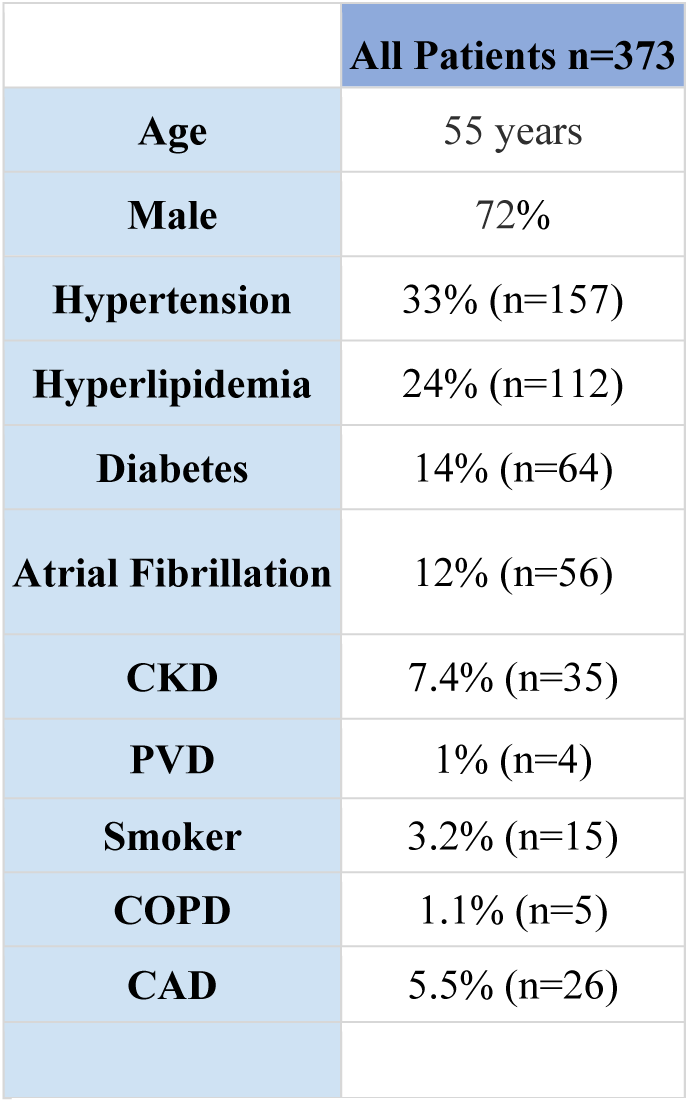
Demographics & Medical History CKD = chronic kidney disease; PVD = peripheral vascular disease; COPD = chronic obstructive pulmonary disease; CAD = coronary artery disease.

**Table 3.** Types of VTA and HF. NICM = non-ischemic cardiomyopathy.

| **Type of Ventricular Arrhythmia (14 Studies Included)** | **% Type of Arrhythmia** | **Type of Heart Failure (15 Studies Included)** | **% of Heart Failure** |
| --- | --- | --- | --- |
| **Monomorphic Ventricular Tachycardia** | 17% (n=72) | **NICM** | 23% (n=80) |
| **Polymorphic Ventricular Tachycardia** | 16.7% (n=70) | **Sarcoid** | 1.7% (n=6) |
| **Pleomorphic** | 9% (n=38) | **Arrhythmogenic Right Ventricular Cardiomyopathy** | 8.8% (n=31) |
| **Long QT Syndrome** | 2.1% (n=9) | **Idiopathic** | 25% (n=89) |
| **Ventricular Fibrillation** | 5% (n=21) | **Ischemic** | 31% (n=108) |
|  |  | **Hypertrophic Cardiomyopathy** | 5.1% (n=18) |
|  |  | **Chagas** | 3.4% (n=12) |
|  |  | **Valvular** | 2.5% (n=9) |

### Changes in ICD Shocks post-surgery

A total of 11 studies reported freedom from ICD shocks at 1 year post CSD yielding a pooled event rate of 69.8% (95% CI, 56.4% – 80.4%, 57% heterogeneity) (I^2^ statistic). A total of 13 studies reported freedom from ICD shocks at 6 months which yielded a pooled event rate of 59.1% (95% CI, 46.9% - 70.4%, 47% I^2^) (Figures 3&4).

**Figure 3.**
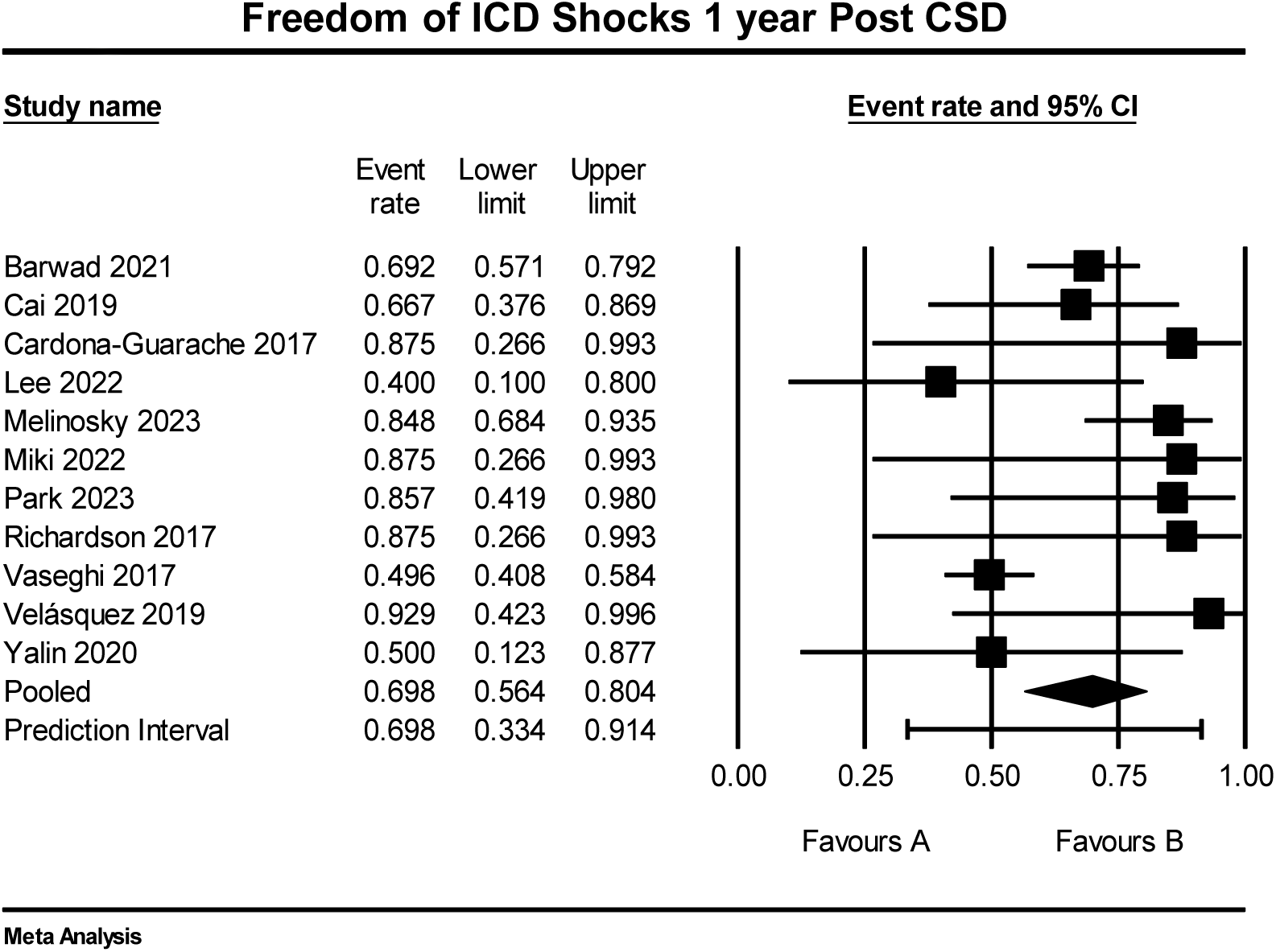
Forest plot event rate of ICD shocks 1 year post CSD where B indicates a ≥ 75% freedom of ICD shocks.

**Figure 4.**
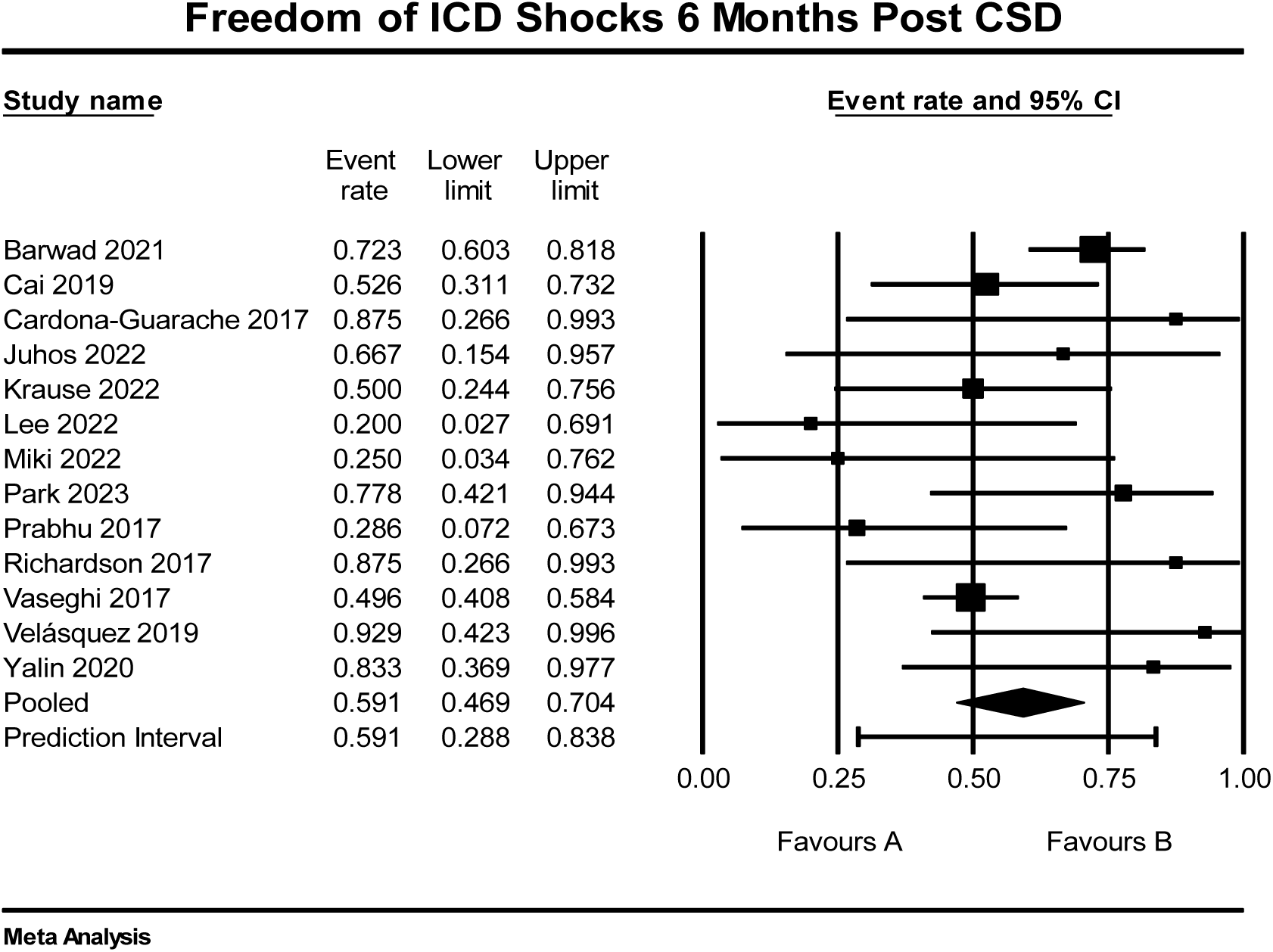
Forest plot showing event rate of ICD shocks at 6 months post CSD where B indicates a ≥ 75% freedom of ICD shocks.

### Changes in VTA post-surgery

A total of 9 studies reported freedom from VTA 1 year post CSD with a pooled event rate of 64.3% (95% CI, 42.3% - 81.5%, 26% I^2^). There were 11 studies that reported freedom from VTA at 6 months with a pooled event rate of 62.3% (95% CI, 51.2% - 72.2%, 40% I^2^) (Figures 5&).

**Figure 5.**
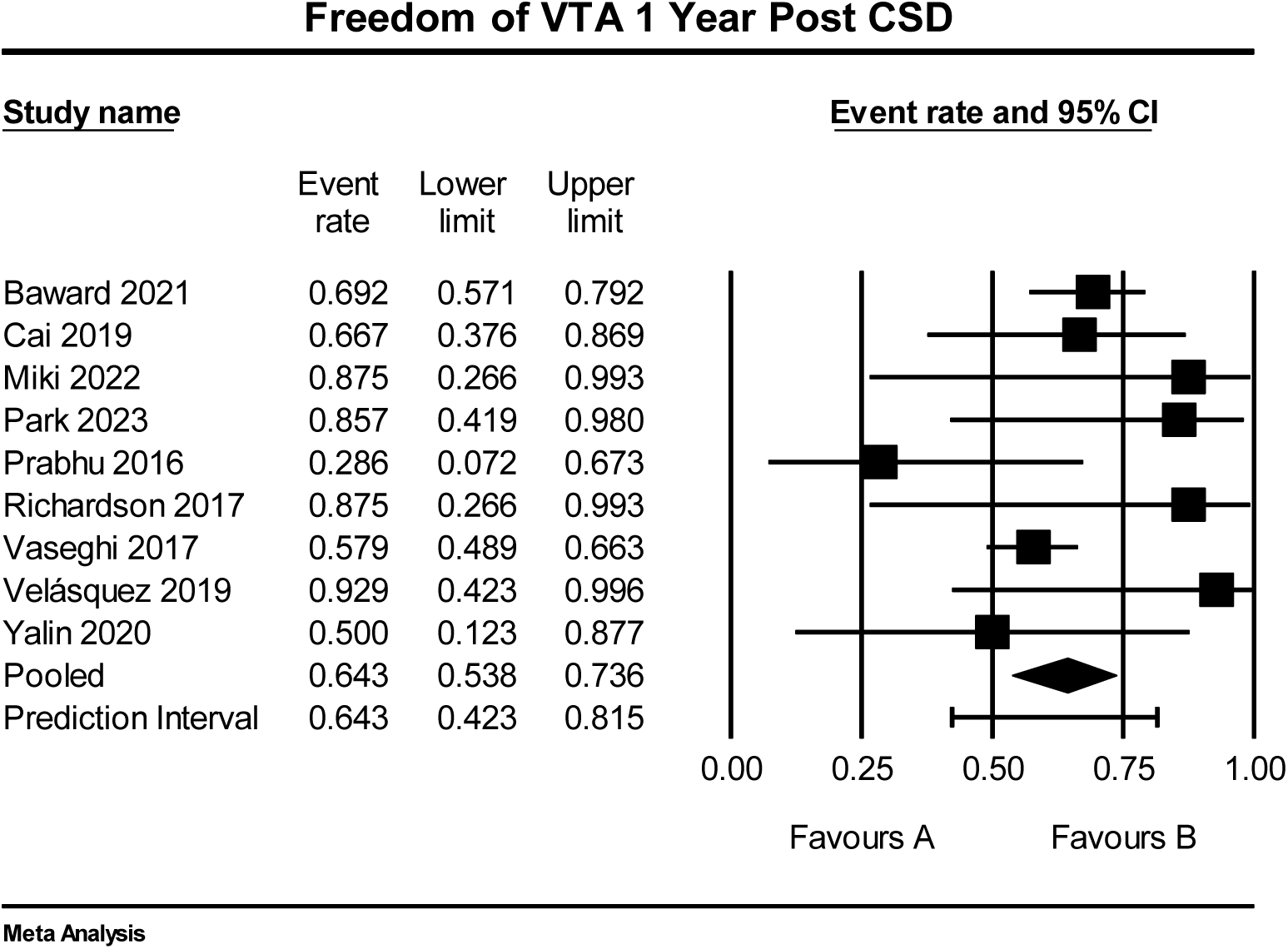
Forest plot of event rate of recurrent ventricular arrhythmias 1 year post CSD where B indicates a ≥ 75% freedom of ICD shocks.

**Figure 6.**
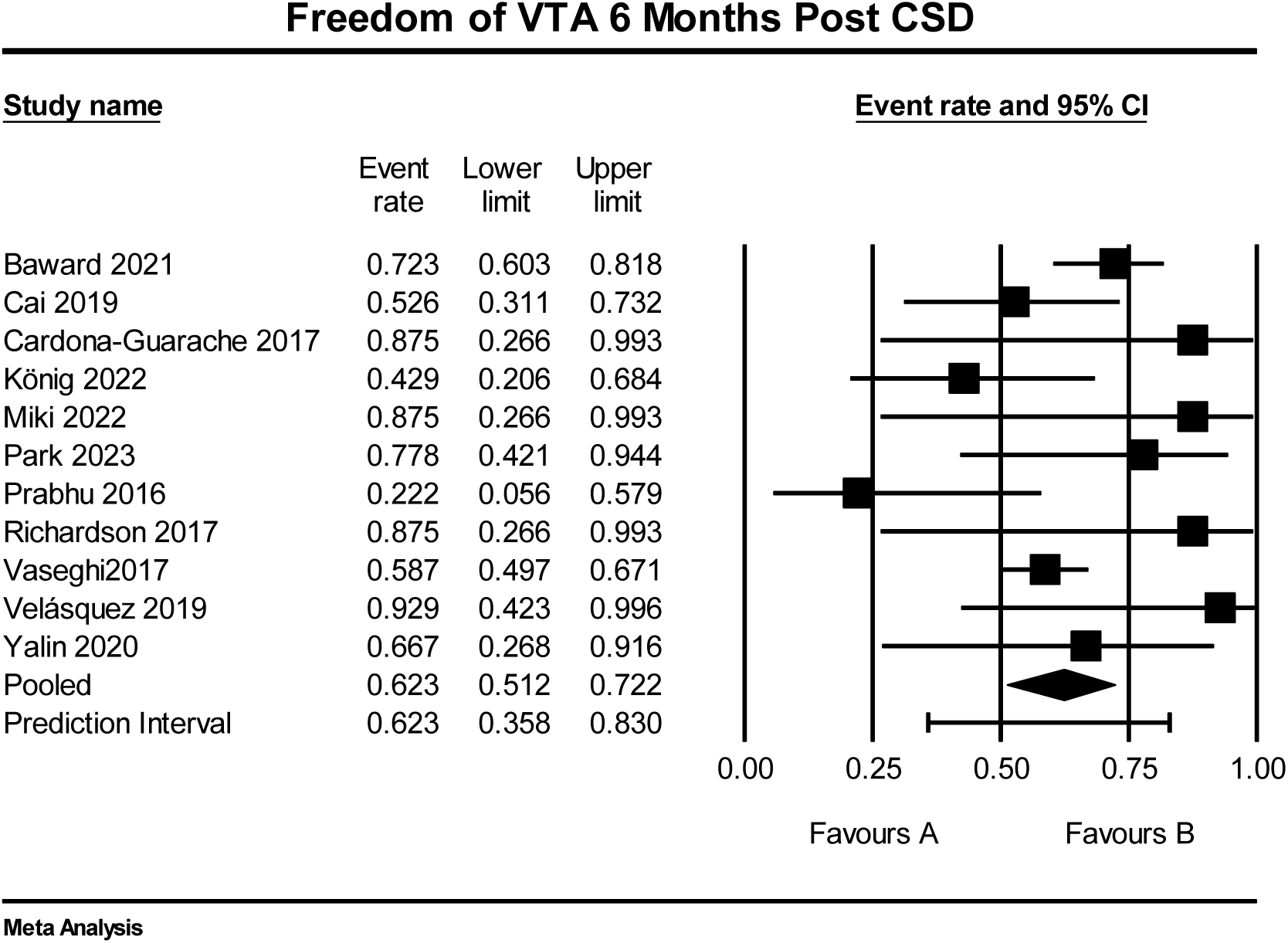
Forest plot of event rate of recurrent ventricular arrhythmias 6 months post CSD where B indicates freedom of ICD shocks ≥75%.

### Temporal Mortality of CSD

Reported mortality due to VTA was subdivided into short-term (0-30 days), intermediate-term (31-364 days) and long-term (>364 days). The pooled event rate of mortality for short-term tertile was 8.9% (95% CI, 5.0% - 15.4%, 0% I^2^), medium term was 5.3% (95% CI, 2.4% - 11.3%, 0% I^2^) and long-term 5.2% (95% CI, 2.4% - 10.9%, 0% I^2^) (Figures 7-9).

**Figure 7.**
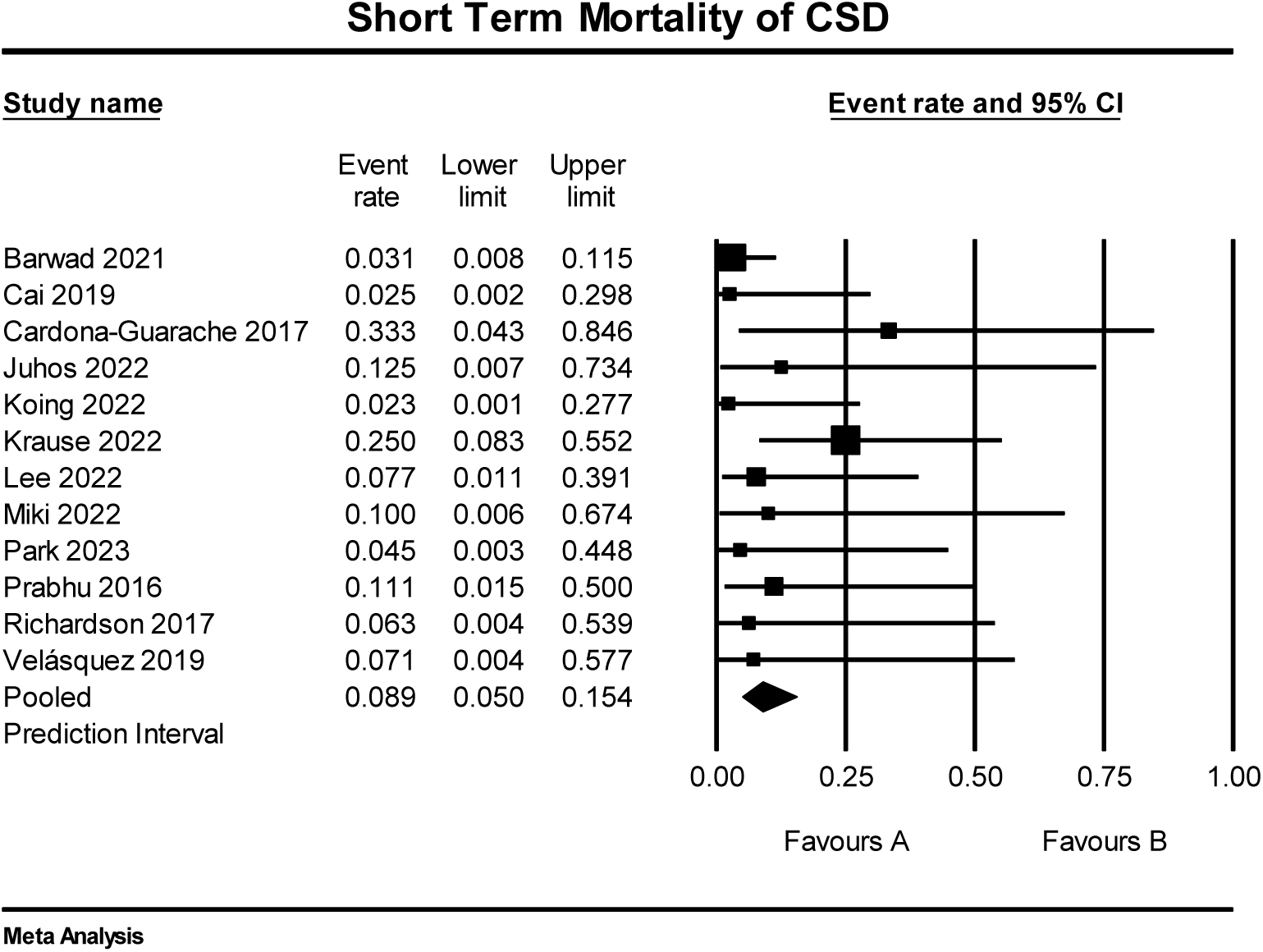
Forrest plot for short term mortality of CSD where A = mortality event rate < 25%.

**Figure 8.**
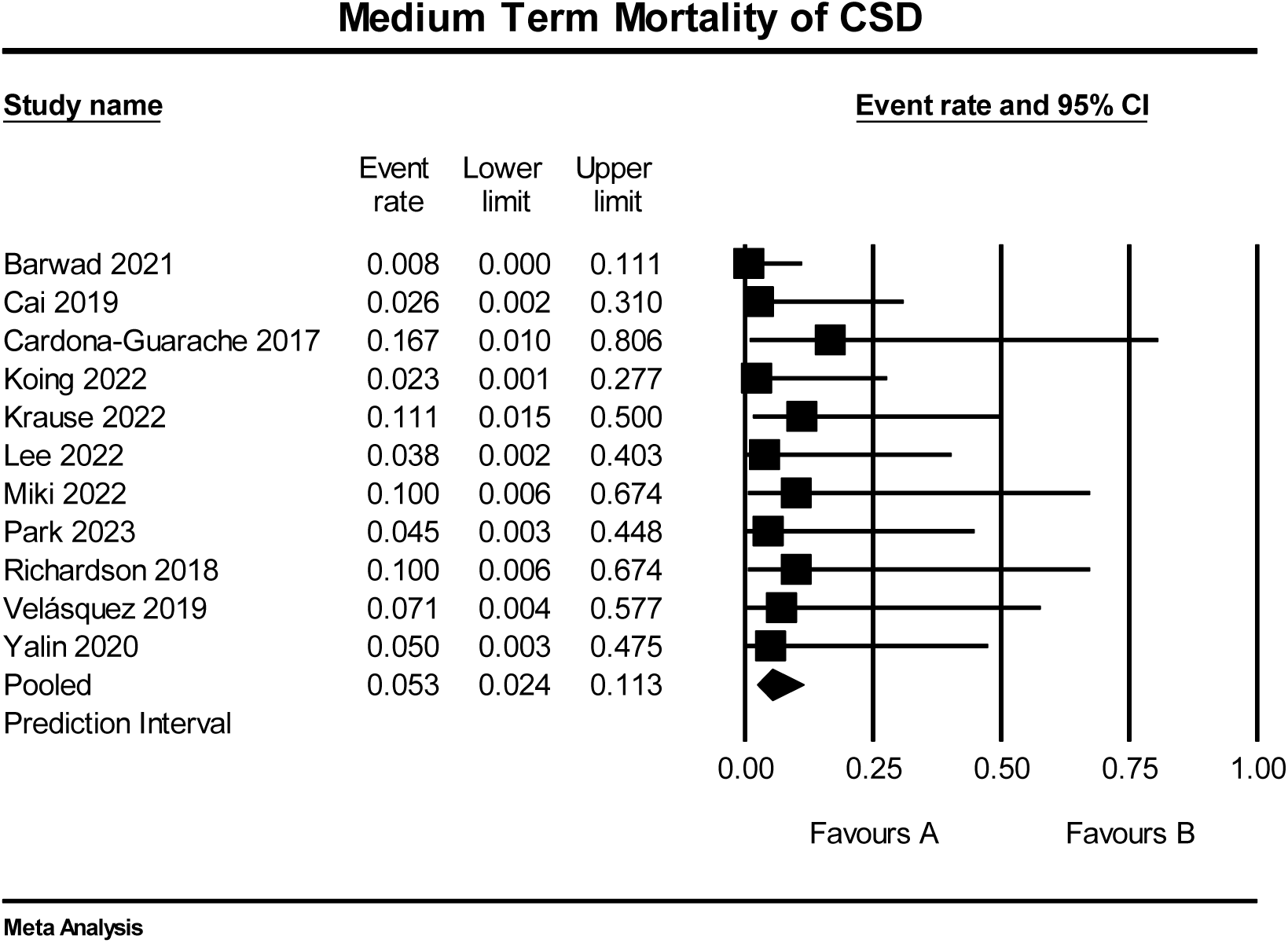
Forrest plot for medium term mortality of CSD where A = mortality event rate < 25%.

**Figure 9.**
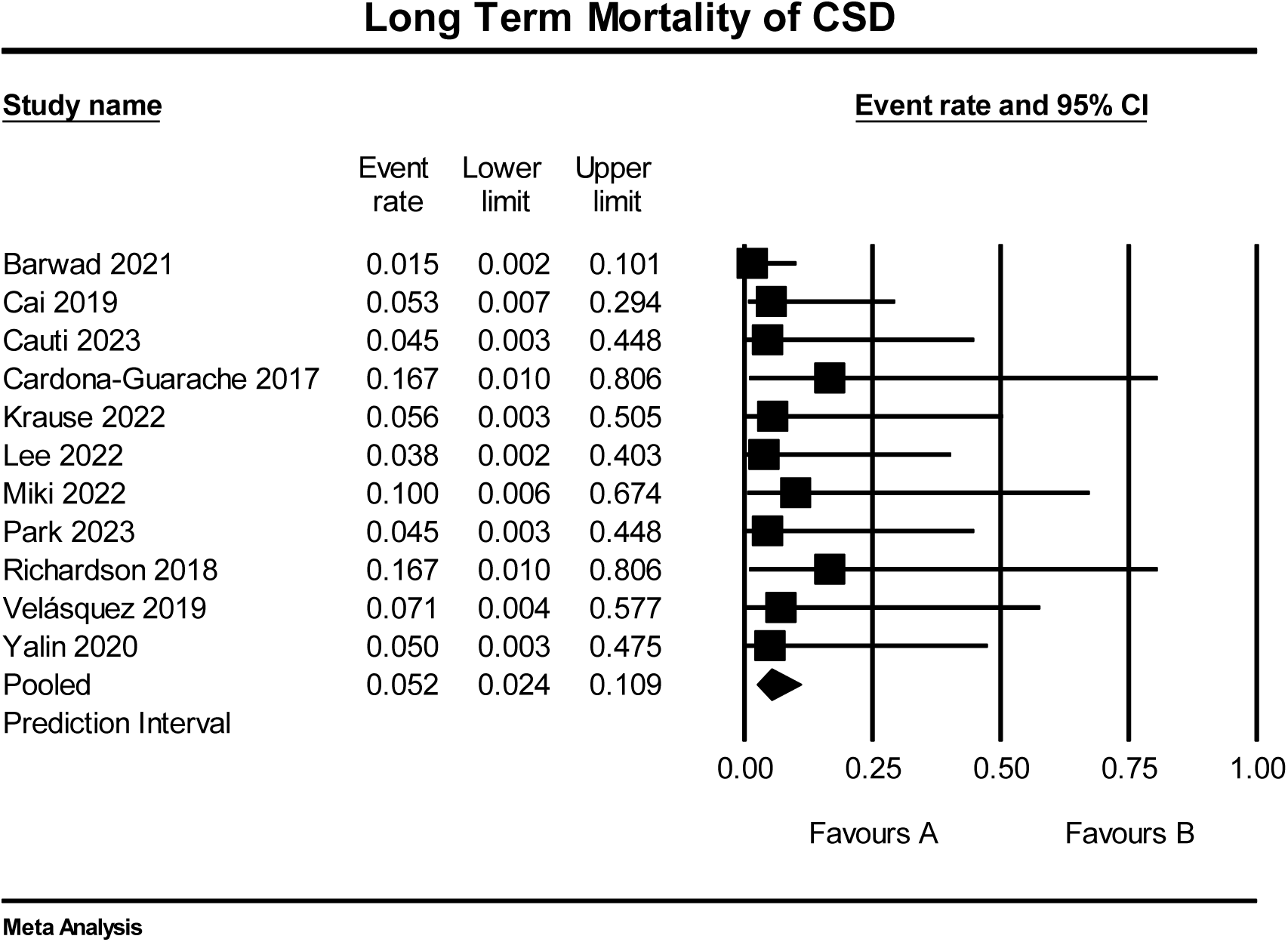
Forrest plot for long term mortality of CSD where A = mortality event rate < 25%.

### Changes in ablation post-surgery

A total of 11 studies provided the percentage of patients that underwent ablation pre and post CSD. On average, 69% of patients underwent cardiac ablation prior to CSD vs. only 15% after CSD (p<0.0003 CI 33.2 80.1).

## Discussion

This contemporary meta-analysis reports the largest series of adult patients suffering from recurrent VTA with structural heart disease who underwent left or bilateral CSD for VTA refractory to antiarrhythmic therapy and/or catheter ablation. Two similar previous meta-analyses have covered this topic.^23,24^ This meta-analysis differs from previous analyses done by Shah et al. and Murtaza et al. by including 3 new studies, only including adults, and reports on incidence of mortality after CSD.

Utilization of CSD was first used in 1961 by Estes et al. in a patient with valvular heart disease and later in 1971 Moss and McDonald in a women experiencing recurrent episodes of ventricular fibrillation due to idiopathic LQTS.^25,26^ The use of CSD was later used by Wilde et al, Collura et al, and De Ferrari et al, in patients with polymorphic ventricular tachycardia.^27–29^ In 1981 Schwartz and Stone verified that left stellectomy applies a noteworthy protective effect in reducing ventricular fibrillation in dogs with myocardial ischemia.^30^ This was eventually expounded further by Schwartz and Vanoli and Schwartz et al by revealing the autonomic mechanism in stellectomys.^31,32^ This eventually led to a clinical trial which revealed the usefulness of surgical and pharmacological antiadrenergic interventions in lowering mortality in patients who are at high risk for sudden cardiac death.^33^ Bourke et al then reported the first reduction of VTA in patients with ischemic cardiomyopathy or non-ischemic cardiomyopathy after left-sided CSD.^34^

Lately, CSD has gained importance as a reasonable substitute in patients with refractory VTA. This meta-analysis demonstrates that patients who undergo CSD benefit from significant VTA and ICD shock reduction by more than 60% at both 6-month and 1-year intervals. This is consistent with prior meta-analyses by Murtaza et al., which reported a pooled rate of freedom from VTA at 60% but with a mean follow-up time of 10 months.^23^ Additionally, this is the first meta-analysis that reports mortality from VTA after CSD to be less than 6% in the short-, medium-, and long-term intervals.

Most of the studies selected patients for CSD who had failed traditional therapy with antiarrhythmic medications and previous cardiac ablations and were still suffering from VTA and VT storms. Most patients underwent multiple failed catheter ablations prior to CSD (Table 1). VT storm was defined as 3 episodes of VTA within 24 hours. Definition of VTA recurrence was based on data from ICD interrogations (any sustained VTA: ventricular fibrillation, monomorphic or polymorphic ventricular tachycardia) or 12-lead ECGs in case of documented sustained VTs below ICD detection.

Multiple studies have shown that patients with cardiac ischemia, inherited arrhythmias, and heart failure predisposes them to have sympathetic activation which in return can cause or worsen VTA.^35,36^ Currently, CSD is a class I recommendation for long QT syndromes and catecholaminergic polymorphic ventricular tachycardia in the 2017 AHA/ACC/HRS guideline in the management of VTA refractory to maximum tolerated beta-adrenergic blockade.^37^ However, much of this evidence is from pediatric studies and patients under the age of 18 years. As previously reported in a combined assessment of the association between VTA burden and mortality, patients who received 1 shock had a fourfold increased risk of death and patients with ≥ 2 shocks had an eightfold increase in death, compared with patients who received no ICD shocks.^37,38^ This favorably translates benefit in this review as CSD lowers rate of ICD shocks. This review also found that the mean number of ICD shocks decreased by 40% when comparing a 1-year pre-CSD to 1 year post CSD. Unfortunately, only 6 studies reported this data and with high degree of heterogeneity, making statistical analysis unfruitful. However, when evaluating the burden of ablation, there was an overwhelming decrease in the percentage of patients requiring repeat ablation post CSD. This is encouraging and might suggest that patients might be demonstrating a better quality of life while exhibiting a reduction in overall symptoms.

Moreover, this review finds that post operative mortality is slightly higher in the first 30 days. The association is unclear, but may be due to comorbidities, end-organ failure in setting of electrical storm, surgical technique, or failure to respond to sympathectomy due to incomplete denervation. Incomplete denervation is linked to the nerve of Kuntz; collateral nerves that bypass the sympathetic trunk form the second to the first intercostal nerve.^23^ Marhold et al. found that detecting the nerve of Kunts was more frequent during open techniques compared to thoracoscopic approach, a technique that is most used in CSD. Failure of complete denervation may result in perpetuation of electrical storm and clinical demise.

In evaluating studies that included both bilateral and unilateral procedures, surgical approach was generally started on the left side followed by the right. Our analysis showed that 79% of our studies conducted bilateral surgical approaches. This was similar to the previous analysis done which found 82% of patients that underwent bilateral carotid sympathetic denervation (BCSD).^24^ Patients who had unilateral CSD and still exhibited VTA, underwent repeat denervation of the opposite sympathetic chain. In these subsets of patients, we found that their response to BCSD was sustained with little no recurrence of VTA or ICD shocks.

Our review found that neuropathic pain and hypotension were the two most common complications with an incidence of 20.5% ± 8.3%. This was similar from previous meta-analysis by Chihara et al. and Shah et al. in which both studies found pneumothorax to be the most common complication. Hypotensive episodes were limited to immediate postoperative period and all patients were successfully weaned off vasopressor medications. Since the procedure involved sympathetic denervation, Horner’s syndrome was a major concern in many of the studies. However, we found that 17% of patients experienced the complication. The lower complication rate might be explained by how surgical technology has advanced with the use of 3-dimensional cameras, and wristed instruments to help with better visualization of the nerve during VATS.

Although this series and others have demonstrated the benefit and risks of CSD in a retrospective observational way, there is quite a paucity of high-quality randomized trial data for this therapy for patients with refractory VTA. It is still unclear as to the appropriate timing to pursue CSD over other traditional standards of care which include further escalation of antiarrhythmic agents, catheter ablation, and cardiac transplantation. Furthermore, the safety of this procedure is still concerning, given that complication rates are high, the long-term implications of chronotropic incompetence after CSD in patients who ultimately undergo cardiac transplantation are unknown. An ongoing randomized clinical trial “Cardiac Sympathetic Denervation for Prevention of Ventricular Tachyarrhythmias (PREVENT VT)” may answer some of these questions.^38^

## Conclusion

In patients with electrical storm and refractory VTA, CSD is a feasible adjunctive therapy to reduce the burden of VTA and ICD shocks once traditional therapies such as antiarrhythmic drug escalation and catheter ablation have been exhausted. The CSD procedure has some risk of complication and morbidity/mortality that needs further investigation.

### Limitations

This systematic review and meta-analysis suffers from limitations and biases of retrospective data collection and analysis. Moreover, meta-analyses have inherent quality issues due to heterogeneity of the data across included studies, poor quality data, selection biases, and large standard errors. There were also no control studies included in this systematic review. Finally, conclusions regarding the efficacy of CSD in comparison to anti-arrhythmic therapy was not possible.

## Data Availability

All data generated or analyzed during this study are included in this published article and its supplementary information files. The datasets generated and analyzed during the current study are available to open access.

## Acknowledgements

None

## Abbreviations

VTA: Ventricular Tachyarrhythmias
CSD: Cardiac sympathetic denervation
HF: heart failure
EF: Ejection Fraction
ICD: Implanted Cardiac Defibrillator

